# Malaria trends in districts that were targeted and not-targeted for seasonal malaria chemoprevention in children under five years of age in Guinea, 2014–2021

**DOI:** 10.1101/2023.09.10.23295324

**Authors:** D Bisanzio, MS Keita, A Camara, T Guilavogui, T Diallo, H Barry, A Preston, L Bangoura, E Mbounga, L Florey, JL Taton, A Fofana, R Reithinger

**Affiliations:** RTI International, Washington D.C., U.S.A.; PMI StopPalu+ Project, RTI International, Conakry, Guinea; National Directorate of Disease Control, Ministry of Health, Conakry, Guinea; National Malaria Control Program, Conakry Guinea; RTI International, Fort Collins, CO, U.S.A.; U.S. President’s Malaria Initiative, US Agency for International Development, Conakry, Guinea; U.S. President’s Malaria Initiative, US Agency for International Development, Washington D.C., U.S.A.

**Author notes:** Corresponding author: Richard Reithinger.

**Keywords:** malaria, seasonal malaria chemoprevention, effectiveness, control, Guinea

## Abstract

**Background:** Seasonal malaria chemoprevention (SMC) is one of the main interventions recommended by WHO to prevent and reduce childhood malaria. Since 2015, Guinea has implemented SMC targeting children aged 3–59 months (CU5) in districts with high and seasonal malaria transmission.

**Objective:** We assessed the programmatic impact of SMC in Guinea’s context of scaled-up malaria intervention programming by comparing malaria-related outcomes in 14 districts that had (n = 8) or had not (n = 6) been targeted for SMC.

**Method:** Using routine health management information system data, we calculated the district-level monthly test positivity rate (TPR) and monthly uncomplicated and severe malaria incidence for the whole population and disaggregated age groups (<5yrs and ≥5yrs of age). Changes in malaria indicators through time were analyzed by calculating the district-level compound annual growth rate (CAGR) from 2014 to 2021; we used statistical analyses to describe the time trend of the number of tested clinical cases, TPR, uncomplicated malaria incidence, and severe malaria incidence.

**Result:** The CAGR of TPR of all age groups was statistically lower in SMC (median = −7.8%, range [IQR] = −9.7%, −5.5%) compared to non-SMC (median = −3.0%, IQR = −3.0%, −1.2%) districts. Similarly, the CAGR in uncomplicated malaria incidence was significantly lower in SMC (median = 1.8%, IQR = −0.9%, 3.5%) compared to non-SMC (median = 11.5%, IQR = 8.8%, 14.0%) districts. For both TPR and uncomplicated malaria incidence the observed difference was also significant age disaggregated. The CAGR of severe malaria incidence showed that all age groups experienced a decline in severe malaria in both SMC and non-SMC districts. However, this decline was significantly higher in SMC (median = −22.3%, IQR= −27.6%, −18.2%) than in non-SMC (median = −5.1%, IQR= - 7.7; −3.6) districts for the entire population, as well as both CU5 and people over 5 years of age.

**Conclusion:** Our results provide evidence to support that—even in an operational programming context—adding SMC to the comprehensive package of malaria interventions yields a positive epidemiological impact and results in greater reduction in TPR, as well as the incidence of uncomplicated and severe malaria in CU5.

## INTRODUCTION

Malaria is the leading cause of morbidity and mortality in Guinea, with 2,422,445 confirmed cases and 1,029 deaths reported in 2021.^1^ Over the past decade Guinea’s National Malaria Control Programme (NMCP)—in collaboration with bilateral, multilateral, and non-governmental partners—has scaled up malaria prevention and control efforts, including rapid diagnostic tests (RDTs), artemisinin-based combination therapies (ACTs), intermittent preventive treatment for pregnant women (IPTp) with sulfadoxine–pyrimethamine (SP), and insecticide-treated nets (ITNs).^2^ Additionally, in 2015, the NMCP piloted and then rolled out seasonal malaria chemoprevention (SMC) in children aged 3-59 months (CU5). Recommended by the WHO since 2012 for countries with seasonal malaria transmission,^3^ SMC is the monthly administration of a single dose of sulfadoxine–pyrimethamine (SP) and three daily doses of amodiaquine (AQ) (SP-AQ) to CU5 during the peak malaria transmission season. SMC can reduce the incidence of clinical malaria in the 28 days following administration by up to 75% when effectively deployed;^4–7^ depending on the length of the rainy season, three to five cycles of SMC are conducted.^8^ As of 2021, 13 countries had adopted SMC (i.e., Benin, Burkina Faso, Cameroon, Chad, Gambia, Ghana, Guinea, Guinea Bissau, Mali, Niger, Nigeria, Senegal and Togo) at different scales of implementation.^8^

Clinical trials and meta-analyses have demonstrated the efficacy of SMC to reduce malaria incidence during the intervention period and parasitemia prevalence at the end of the transmission season, and suggest a positive impact on all-cause mortality.^4–7^ There is also some evidence that SMC results in lower prevalence of moderate to severe anemia and is associated with greater weight gain or improvement of malnutrition indicators, but such effects have not always been significant across studies.^7, 9–14^ Monthly administration of SP-AQ was found to be the drug regimen with the highest efficacy,^15–17^ and using community health workers (CHWs) to deliver SMC was shown to be more cost-effective than using facility-based nurses, immunization outreach clinics, or outreach trekking teams.^18,19^

While there is strong evidence of the effect of SMC in rigorously conducted academic research studies, evidence on the protective effect of SMC in programmatic contexts is more limited.^7, 20, 21^ This type of evaluation is challenging but very necessary—indeed, the effectiveness of interventions in the context of public health programs often differs from the efficacy measured in academic research studies, because of operational challenges when interventions are implemented at scale.^22, 23^

The aim of the analyses presented here was to assess the programmatic impact of SMC in Guinea’s context of scaled-up malaria intervention programming, specifically by comparing malaria-related outcomes in districts that had or had not been targeted for SMC between 2015 and 2021. Additionally, to do so, we wanted to use a methodological (i.e., using data routinely collected by Guinea’s health management information system) and an analytical approach (i.e., using the compound annual growth rate) that is simple and would allow the NMCP to readily monitor SMC effectiveness against standard malariometric indicators as it continues to expand the intervention across the country.

## METHODOLOGY

### Study Setting

The Republic of Guinea is located in West Africa: it covers a total land area of 245,860 km^2^ and has an estimated population of 13.5 million people. The country is divided into 8 major administrative regions, which are further divided into 38 préfectures (districts), five of which comprise the urban areas around the capital city, Conakry. Districts are the main administrative unit where many of the public services are planned, managed, and implemented, including for health and malaria. Guinea’s climate is tropical and humid with a wet (June to November) and a dry season (December to May); the rainy season is followed by a peak malaria transmission season, with the highest malaria case count typically observed between July to November / December. Approximately 95% of malaria cases in Guinea are caused by *Plasmodium falciparum*, the principal vectors being *Anopheles gambiae*, *Anopheles funestus*, and *Anopheles arabiensis*.^1, 24, 25^

### SMC in Guinea

Guinea introduced SMC initially in 6 districts in 2015, scaling up to 17 districts by 2021, with four monthly community-based SMC campaigns (cycles) conducted during the peak malaria transmission season (generally from July to October). The NMCP implements SMC in partnership with nongovernmental and international organizations that financially support the National Malaria Strategic Plan and its activities. As per WHO guidance,^8^ the recommended drug regimen for SMC is SP-AQ. CHWs (existing ones as well as ones mobilized specifically for the SMC campaign) administer SP and the first dose of AQ to eligible children and give the remaining two daily doses of AQ to the caregiver.

Cycles of SMC are conducted each year—once every 4 weeks during peak malaria transmission season. SMC usually starts late July, but the exact starting date fluctuates every year depending on several factors, including logistical considerations and rainfall. The target population for SMC comprises all CU5, excluding those with known allergies to SP or AQ, those under cotrimoxazole treatment, and those severely ill or experiencing a presumptive malaria episode. Children 3–11 months of age receive one 25 mg dose of sulphadoxine, one 12.5 mg dose of pyrimethamine, and three doses of 75 mg AQ given over the course of three consecutive days; children 12–59 months of age receive double doses of SP and AQ. The children’s vaccination booklets are consulted to help determine their age.

In teams of two, CHWs go door-to-door to every household; sometimes they organize distribution sessions at gathering venues such as markets, churches, dwellings, mosques, and fields. After explaining the SMC strategy (objective, rationale, risks, and benefits, treatment instructions) to the caregiver, they administer treatments, with any children with malaria or danger signs referred to the closest health centre. CHWs fill in forms where they indicate the number of treatments administered in every household, and they keep updated stock management sheets. Drugs are supplied through the Ministry of Health (MOH) to the health centres, who then allocate them to the CHWs. Nurses in health centres are responsible for coordinating CHWs’ work and for collecting SMC forms. Supervisory visits are conducted by nurses and district health authorities.

### Malaria Data

In the aftermath of the 2014–2016 Ebola outbreak, the MOH established a strategic plan to strengthen its routine surveillance system, which led to the adoption of the District Health Information Software (DHIS) 2, an open source, online software application as the national health information system platform for monthly aggregate data from health facilities. Today this health management information system (HMIS) is managed by Guinea’s *Système National d’Information Sanitaire* (SNIS) and has been rolled out nationally;^26^ a separate instance is also currently used for weekly aggregate disease surveillance and individual case surveillance of epidemic-prone diseases. The NMCP uses DHIS2 to store the monthly aggregate epidemiological surveillance data collected, including all out -and inpatients seen, suspected clinical malaria cases seen, suspected clinical malaria cases tested, malaria cases confirmed, and malaria cases treated.^26^ Monthly numbers of tested clinical malaria cases, confirmed positive clinical cases, and severe malaria are disaggregated by age group (<5 years of age [<5yrs], ≥5 years of ages [≥5yrs], and pregnant women). Data for CU5 and people ≥5 yrs of age (PO5) were downloaded for January 2014 to November 2021 from the SNIS portal; we also downloaded the estimated catchment population of each health facility for each year.

### Study area, Study Population, and Data Analysis

Analyses included 8 (Labé, Koubia, Tougue, Mali, Lelouma, Gaoual, Koundara, and Dinguiraye) and 6 (Boffa, Boké, Coyah, Dubreka, Forecariyah, and Fria) districts where SMC campaigns were or were not performed from January 2014 to November 2021, respectively. We adjusted analyses to account for different year of enrolment of districts in the SMC campaign (i.e., Labé and Lelouma started SMC in 2017). The 14 study districts were among the districts supported by StopPalu and StopPalu+, projects funded by the U.S. President’s Malaria initiative and implemented by RTI International. The remaining districts covered by these two projects were the communes of the capital Conakry (i.e., Matam, Dixinn, Ratoma, Matoto, and Kaloum), which were not eligible for SMC due to their low malaria prevalence; these districts were therefore excluded from the analyses.

Among all the study districts’ facilities reporting data to SNIS, we selected those that had a high level of malaria data reporting completeness. Given the long study period and possible difficulties in consistent data reporting, an arbitrary threshold of 10 missing months within the 2014– 2021 study period and no more than 3 continuously missing months per year was used to identify facilities with adequate data reporting completeness. We only included public health facilities (i.e., hospitals, all health centre types, and health posts) supported by StopPalu and StopPalu+. This selection method identified 131 facilities of the 149 in the dataset with adequate reporting completeness. The resulting catchment population based on the selected facilities was 4,451,792 people in 2014, increasing to 5,244,844 people in 2021.

Using the extracted indicator data from SNIS, we calculated the monthly test positivity rate (TPR) and monthly uncomplicated and severe malaria incidence at the district level for the whole population, as well as disaggregated by age groups (<5yrs and ≥5yrs of age). We used statistical analyses to describe the time trend of the number of tested fevers, TPR, uncomplicated malaria incidence, and severe malaria incidence. The changes of these malaria indicators through time were analyzed by calculating the compound annual growth rate (CAGR) from 2014 to 2021 at the district level. The CAGR was calculated by dividing a time series end value by its beginning value and raising the resulting figure to the inverse number of the time series years subtracting it by one. Thus, for our SMC analyses, the CAGR for each malaria indicator was calculated as follows:

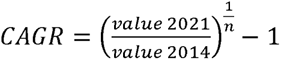

where n is the year of SMC implementation. Testing for differences in malaria indicator values and their CAGR between SMC and non-SMC districts were performed by using the Wilcoxon’s signed rank test.^27^

### Ethical approval

This study only used existing (secondary) data that were collected for public health planning and programming purposes. The data were only available and analyzed at the aggregate level and thus ethical approval for the analyses was not necessary a priori.

## RESULTS

### SMC Coverage

In the districts included in the analyses, 8.1 million treatment doses of SP+AQ were administered to eligible CU5 between 2015 and 2021, resulting in an average annual programmatic SMC coverage of 89% (range between years: 86–93%).

### Number of Clinical Malaria cases Tested

From January 2014 to November 2021, 5,002,551 clinical malaria cases were tested in the health facilities included in the study, of which 1,658,637 were CU5 (33.2%) (**Table 1**). Among all tested clinical cases, 2,484,794 (49.7%) and 2,517,757 (50.3%) were tested in SMC and non-SMC districts, respectively; in both district groups the proportion of clinical malaria in CU5 was approximately 30% of all tested clinical cases (SMC districts = 29.8% *vs* non-SMC districts = 36.5%) (**Table 1**). From 2014 to 2021, the trend of tested clinical cases in CU5 and PO5 showed consistent increase in all districts (**Figure 2**). The 2014–2021 CAGR of tested clinical cases for SMC districts (median = 12.2%, range = 4.1%, 20.6%) was significantly lower compared to that of non-SMC districts (median = 19.3%; range = 12.9%, 22.2 %) (Wilcoxon’s test, p<0.05) (**Figure 3**). When disaggregated by age, the CAGR of tested clinical cases was lower in SMC districts compared to non-SMC districts for both CU5 and PO5, but it was only statistically significant for CU5 (Wilcoxon’s test, p<0.05) (**Figure 3**).

**Figure 1.**
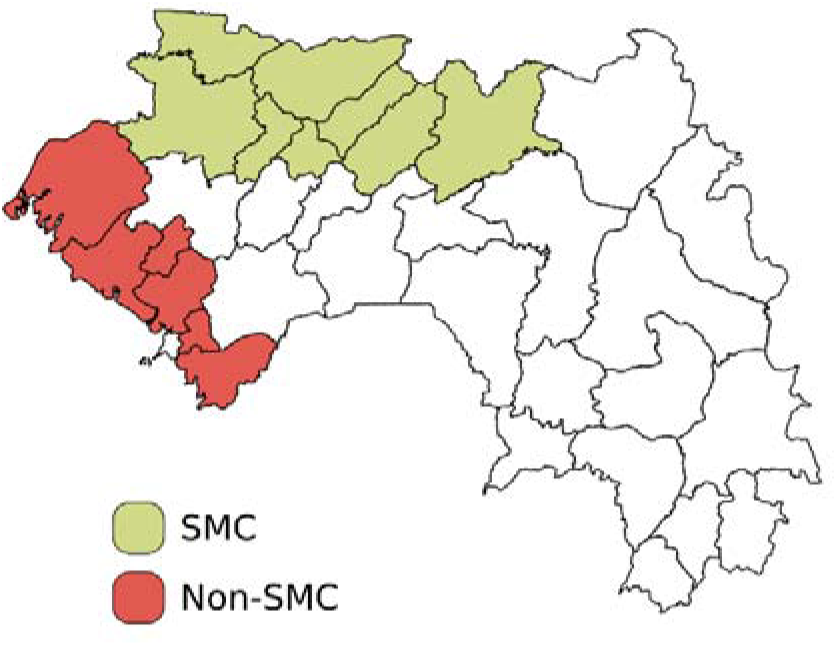
Districts covered by the StopPalu and StopPalu+ in which SMC was performed.

**Figure 2.**
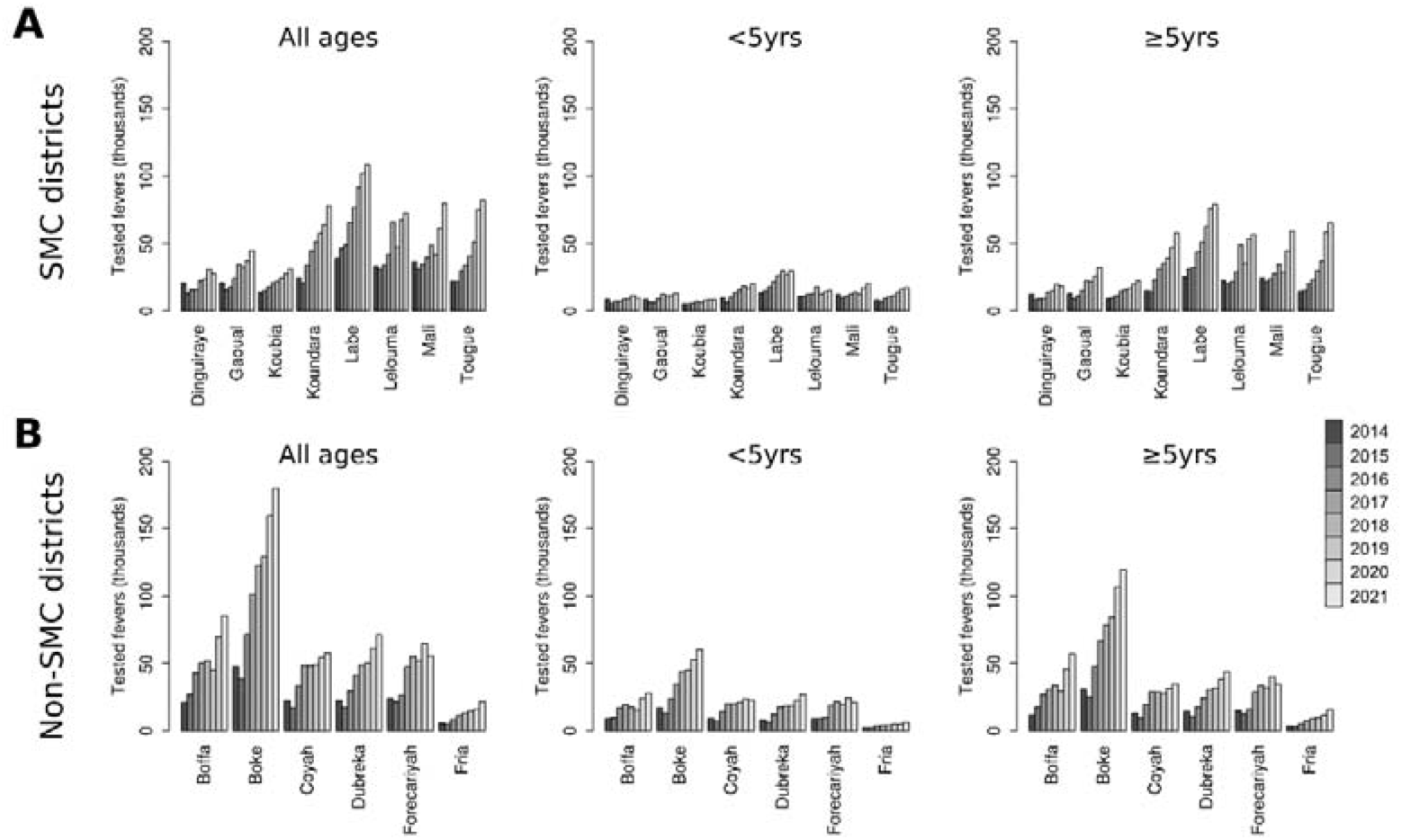
Number of tested fevers in SMC and non-SMC districts per age group and year. Plots show tested fevers for all-age groups, <5yrs age group, and ≥5yrs age group in SMC (A) and non-SMC districts (B)

**Figure 3.**
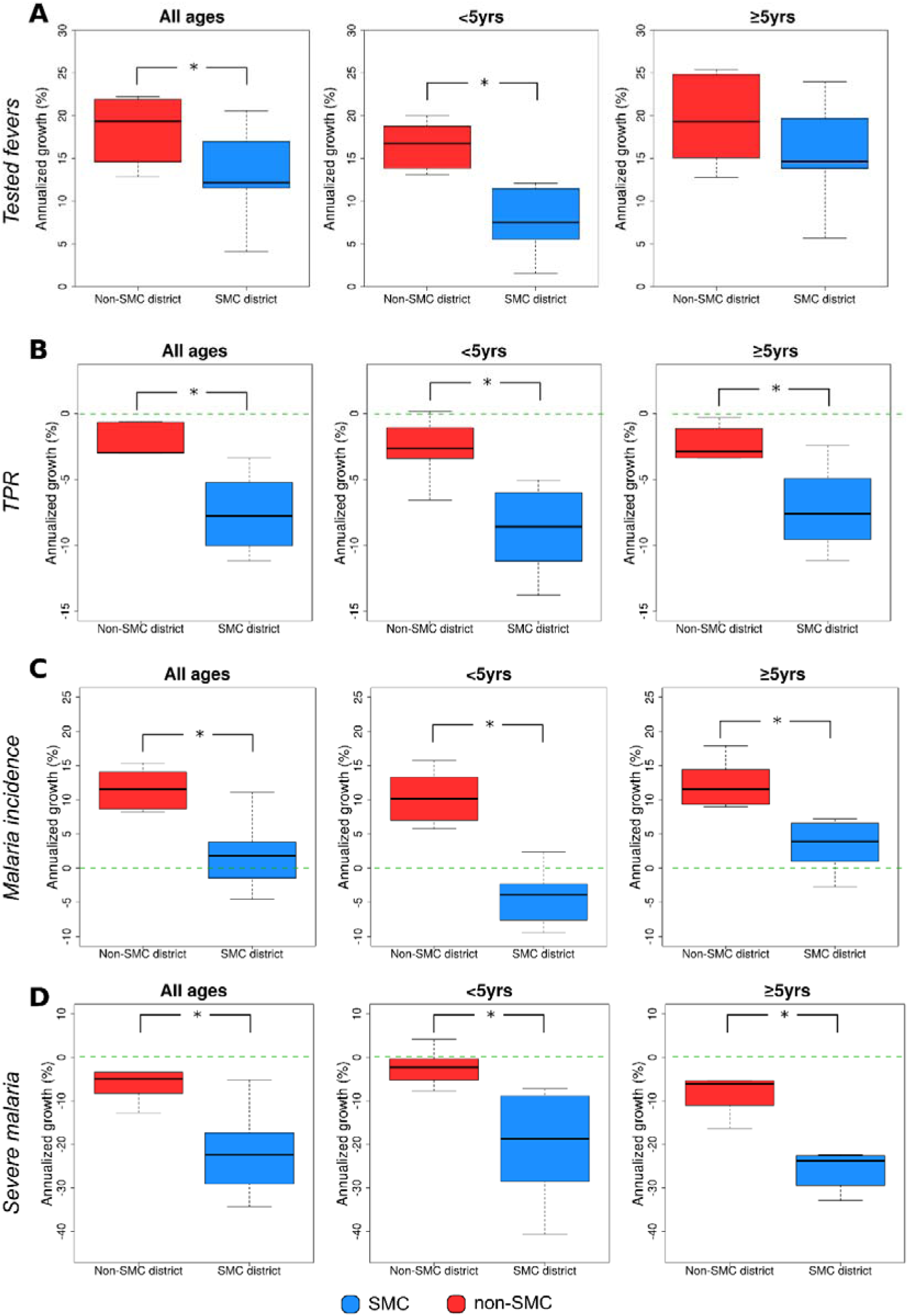
CAGR of tested fevers (A), TPR (B), malaria incidence (C), and severe malaria cases (D) in SMC districts and non-SMC districts from 2014 to 2021. The ‘*’ symbol represents a significant difference (p<0.05, Wilcoxon’s test) between CAGR values of SMC district and non-SMC districts.

**Table 1.** Number of tested fevers, positive tests, and positive rate divided by age group and SMC coverage during the study period (January 2014 – November 2021).

| Age Group | All districts |  | SMC districts |  | Non-SMC district |  |
| --- | --- | --- | --- | --- | --- | --- |
|  | Tested | Positive (%) | Tested | Positive (%) | Tested | Positive (%) |
| <5yrs age group | 1,658,637 | 915,831 (55.2%) | 739,403 | 344,420 (46.6%) | 919,234 | 571,411 (62.2%) |
| ≥5yrs age group | 3,343,914 | 1,928,125 (57.7%) | 1,745,391 | 904,818 (51.8%) | 1,598,523 | 1,023,307 (64.1%) |
| Total | 5,002,551 | 2,843,956 (56.9%) | 2,484,794 | 1,249,238 (50.3%) | 2,517,757 | 1,594,718 (63.4%) |

### Test Positivity Rate

The TPR among individuals who were tested from 2014–2021 was 56.9% (2,843,956 people), with PO5 having a slightly higher TPR (57.7%) compared to CU5 (55.2%) (**Table 1**). When SMC was accounted for, SMC districts had a lower TPR compared to non-SMC districts for each age group, with the difference in TPR being 15.6 and 12.3 percentage points for CU5 and PO5, respectively (**Table 1**). Comparing the TPR among districts from 2014 to 2021, the TPR steadily declined in SMC districts, while the decline in most non-SMC districts stopped in 2019, followed by an increase during 2020 and 2021 (**Figure 4**). The TPR trend during the 2014–2021 study period showed a malaria season characterized by a high transmission season between July and December (Guinea’s rainy season: June–November), followed by a low transmission period occurring between January and May (Guinea’s dry season: December–May) (**Figure 5**); peak malaria transmission usually occurred between August and September. The TPR of SMC and non-SMC districts was similar in 2014 (Wilcoxon’s test, p>0.05), but evolved annually to become significantly different in 2021 (Wilcoxon’s test, p<0.05) (**Figure 5**). When disaggregated by age, for both age groups, the reduction in TPR between 2014 and 2021 in SMC districts (CU5 tested fevers: median = −46.6%, range = −64.6%, - 30.3%; PO5 tested fevers: median = −42.6%, range = −56.2%, −15.8%) was more than two times higher compared to the reduction in non-SMC districts (CU5 tested fevers: median = −17.1%, range = - 37.9%, 1.1%; PO5 tested fevers: median = −18.5%, range = −56.2%, −2.8%) (**Figure 6**). The CAGR of TPR of all age groups was statistically lower in the SMC (median = −7.8%, range = −9.7%, −5.5%) compared to non-SMC (median = −3.0%, range = −3.0%, −1.2%) districts (Wilcoxon’s test, p<0.05, **Figure 3**). When disaggregated by age, the CAGR was significantly different for both CU5 (SMC districts: median = −8.6%, range = −10.9%, −6.3%; non-SMC districts: median = −2.6%, range = −3.3%, −1.4%) and PO5 (SMC districts: median = −7.6%, range = −9.4%, −5.2%; non-SMC districts: median = −2.9%, range = −3.3%, −1.5%) when comparing the two district groups (Wilcoxon’s test, p<0.05, Figure 3).

**Figure 4.**
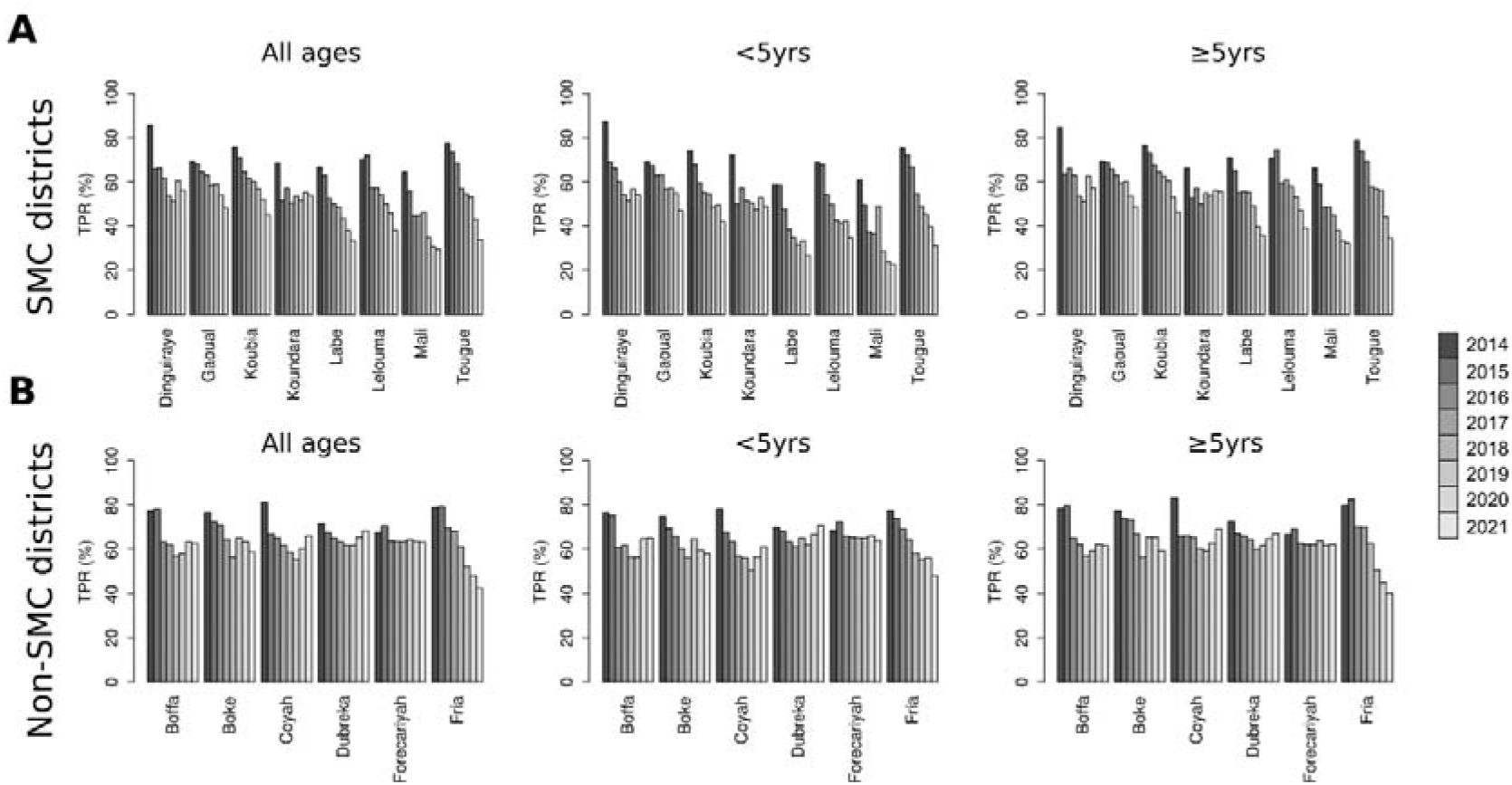
TPR of tested fevers in SMC districts and non-SMC districts per age group and year. Plots show TPR for all-age groups, <5yrs age group, and ≥5yrs age group in SMC (A) and non-SMC districts (B)

**Figure 5.**
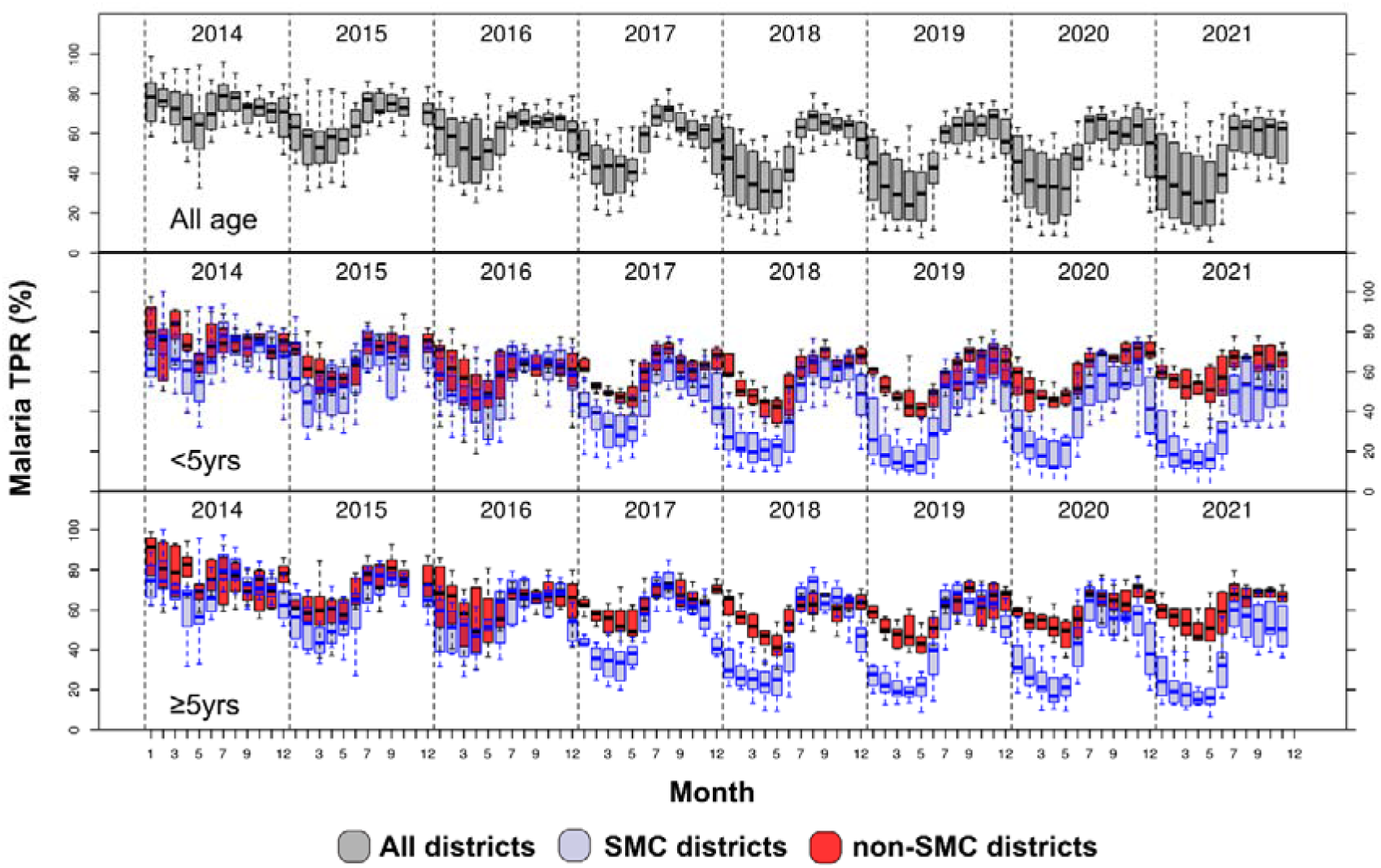
Boxplot of annual TPR of SMC districts and non-SMC districts from January 2014 to November 2021 for all age groups, <5 age group, and **≥**5 age group.

**Figure 6.**
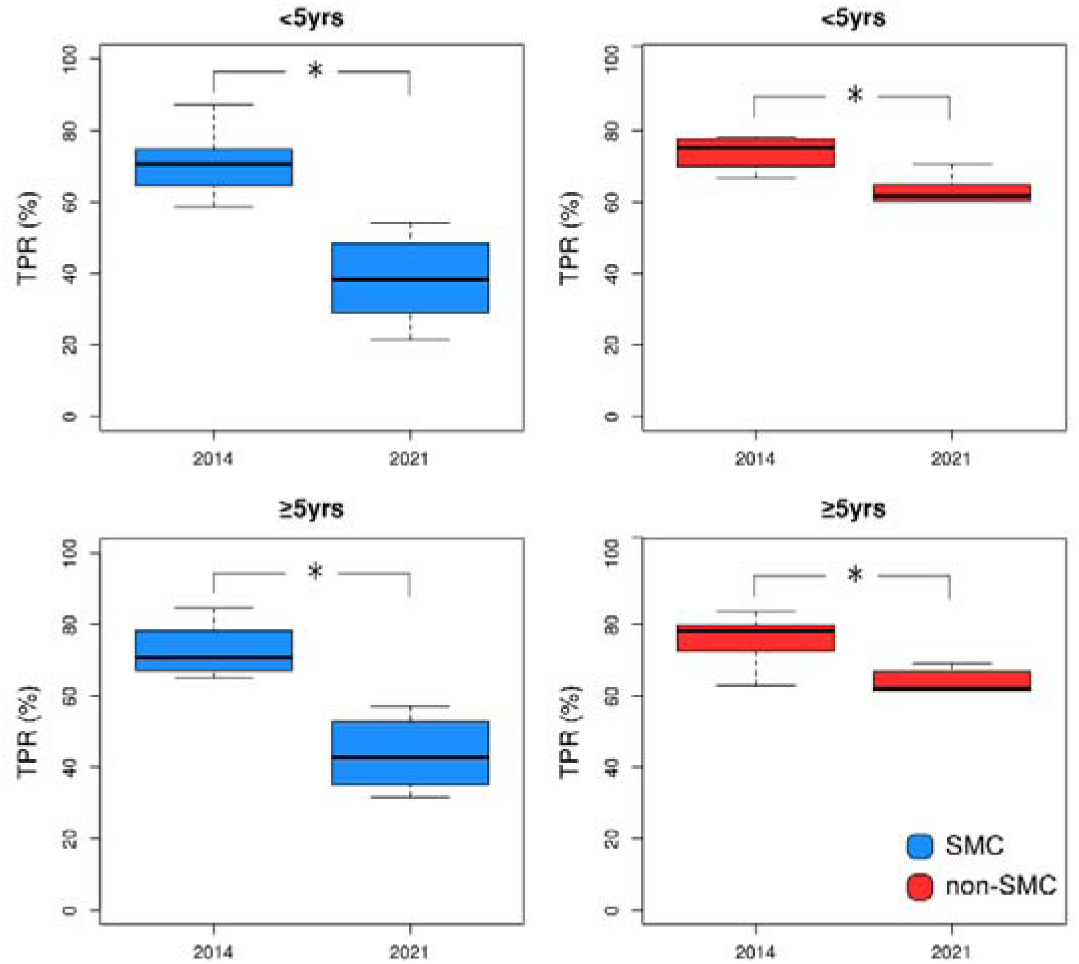
Boxplot comparing TPR of SMC (left column) and non-SMC (right column) districts by age group of 2014 to November 2021 for <5 yrs and ≥5 yrs age group. The ‘*’ symbol represents a significant difference (p<0.05, Wilcoxon’s test) between SMC and non-SMC districts.

### Uncomplicated Malaria Incidence

The median malaria incidence in SMC districts was 18.5 cases per 1,000 people (IQR = 14.5, 21.6) in 2014 and 19.1 cases per 1,000 people (IQR = 16.4, 28.6) in 2021 (**Figure 7**). The median incidence in non-SMC districts was 10.3 cases per 1,000 people (IQR = 9.1, 17.3) in 2014 and 26.7 cases per 1,000 people (IQR = 18.2, 33.7) in 2021. Thus, both SMC and non-SMC districts experienced an increase of incidence from 2014 to 2021. Comparing the incidence trend among the age groups and districts, only the incidence of CU5 in SMC districts showed a declining trend from 2014 to 2021. Incidence showed to be statistically lower during the low transmission seasons in the SMC districts compared to non-SMC districts (Wilcoxon’s test) (**Figure 8**). The CAGR in all-age malaria incidence in SMC districts (median = 1.8%, range = −0.9%, 3.5%) was significantly lower compared to non-SMC districts (median = 11.5%, range = 8.8%, 14.0%) (Wilcoxon’s test p<0.05, **Figure 3**). When disaggregated by age, the CAGR between SMC and non-SMC districts were significantly different for both CU5 (SMC districts: median = −3.9%, range = −7.6%, −2.6%; non-SMC districts: median = 10.2%, range = 7.5%, - 12.9%) and PO5 (SMC districts: median = 3.9%, range = 1.6%, 6.3%; non-SMC districts: median = 11.5%, range = 9.6%, 14%) when comparing the two district groups (Wilcoxon’s test, p<0.05, **Figure 3**).

**Figure 7.**
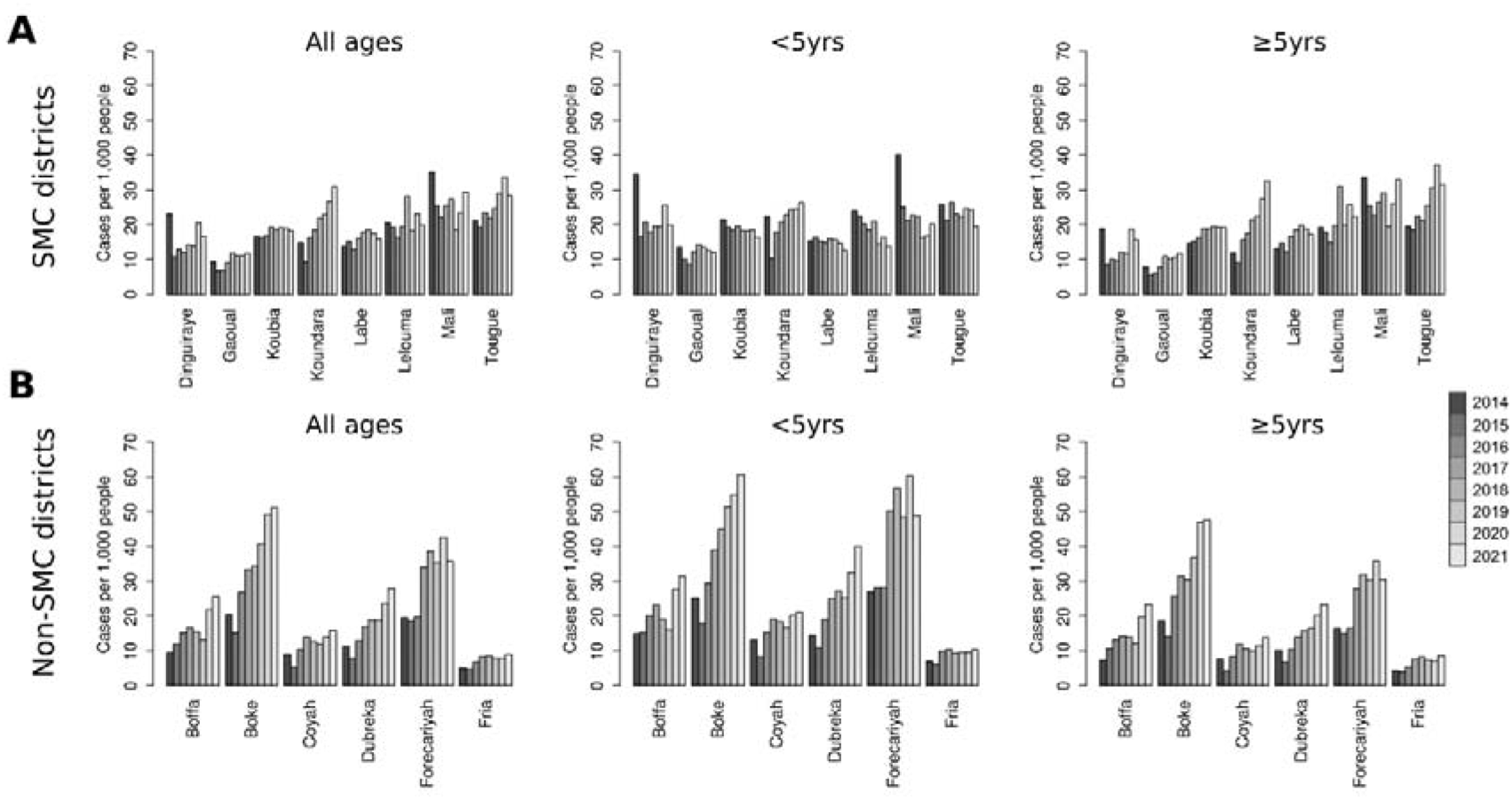
Malaria incidence in SMC (A) and non-SMC (B) districts per age group and year. Plots show malaria incidence for all-age groups, <5yrs age group, and ≥5yrs age group in SMC (A) and non-SMC districts (B)

**Figure 8.**
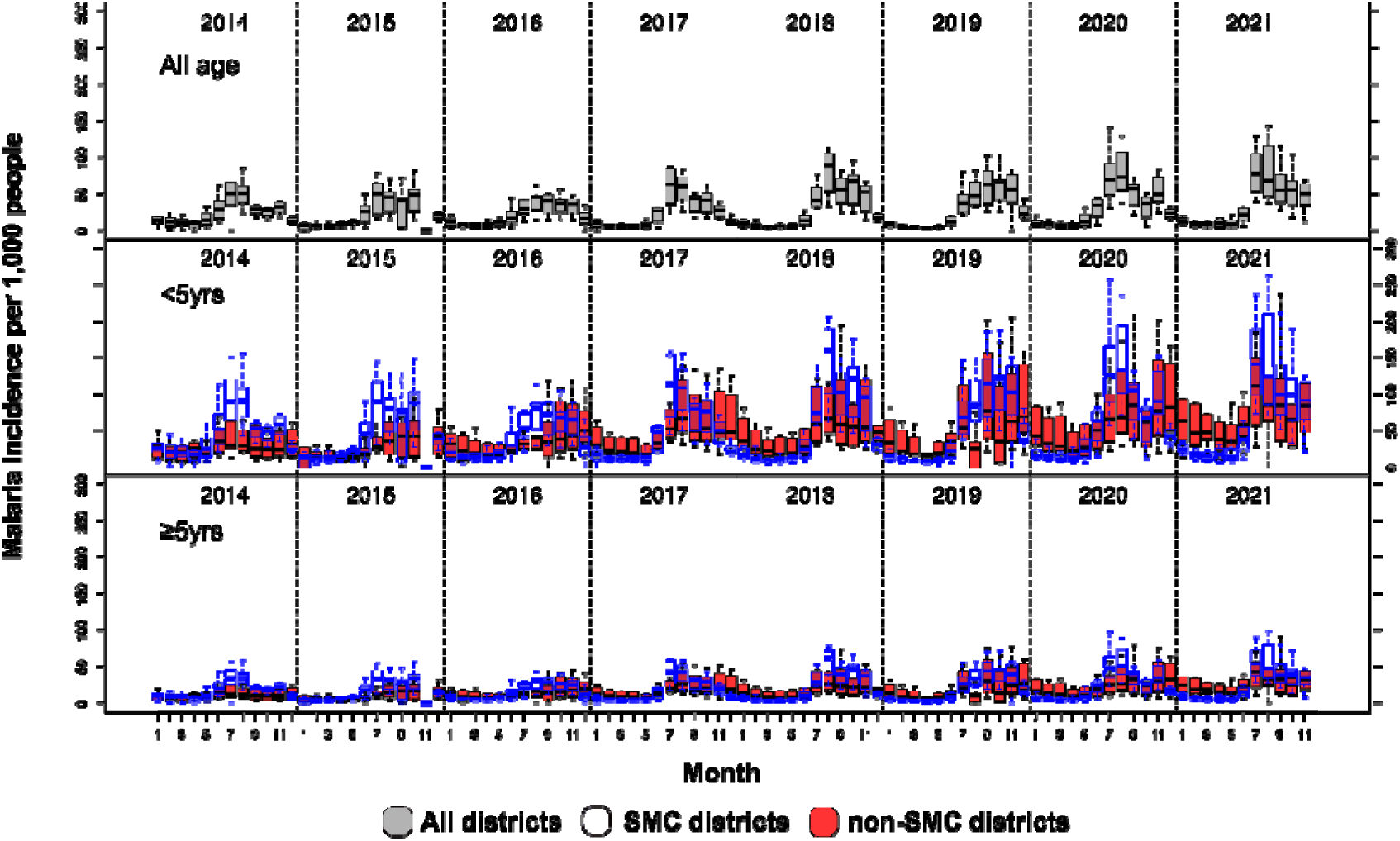
Boxplot of malaria incidence of SMC districts and non-SMC districts from January 2014 to November 2021 for all age groups, <5 yrs age group, and ≥5 yrs age group, by year.

### Number of Severe Malaria Cases

From January 2014 to November 2021, 264,726 severe malaria cases were reported from the facilities included in the study, of which 111,647 (42.2%) were from SMC districts and 153,079 (57.8%) from non-SMC districts. The proportion of severe malaria in CU5 was similar in SMC districts (40,426 cases, 36.2%) and non-SMC districts (56,459 cases, 36.8%). The median severe malaria incidence in SMC districts declined from 1.4 cases per 1,000 people (IQR = 1.1, 1.6) in 2014 to 0.2 cases per 1,000 people (IQR = 0.1, 0.3) in 2021, and from 0.9 cases per 1,000 people (IQR = 0.8, 1.1) to 0.6 cases per 1,000 people (IQR = 0.5, 0.7) in non-SMC districts (**Figure 9**). The incidence of severe cases was higher in PO5 in both SMC districts and non-SMC districts. The reduction of severe malaria cases declined in both age groups, but was significantly higher in the CU5 (Wilcoxon’s test p<0.05) (**Figure 10**). The CAGR of severe malaria incidence showed that all age groups experienced a decline in severe malaria in both SMC and non-SMC districts. However, this decline was significantly higher in SMC (median = −22.3%, range = −27.6%, −18.2%) than in non-SMC (median = −5.1%, range = −7.7, - 3.6) districts for the entire population, as well as both CU5 and PO5 (Wilcoxon’s test p<0.05) (**Figure 3**).

**Figure 9.**
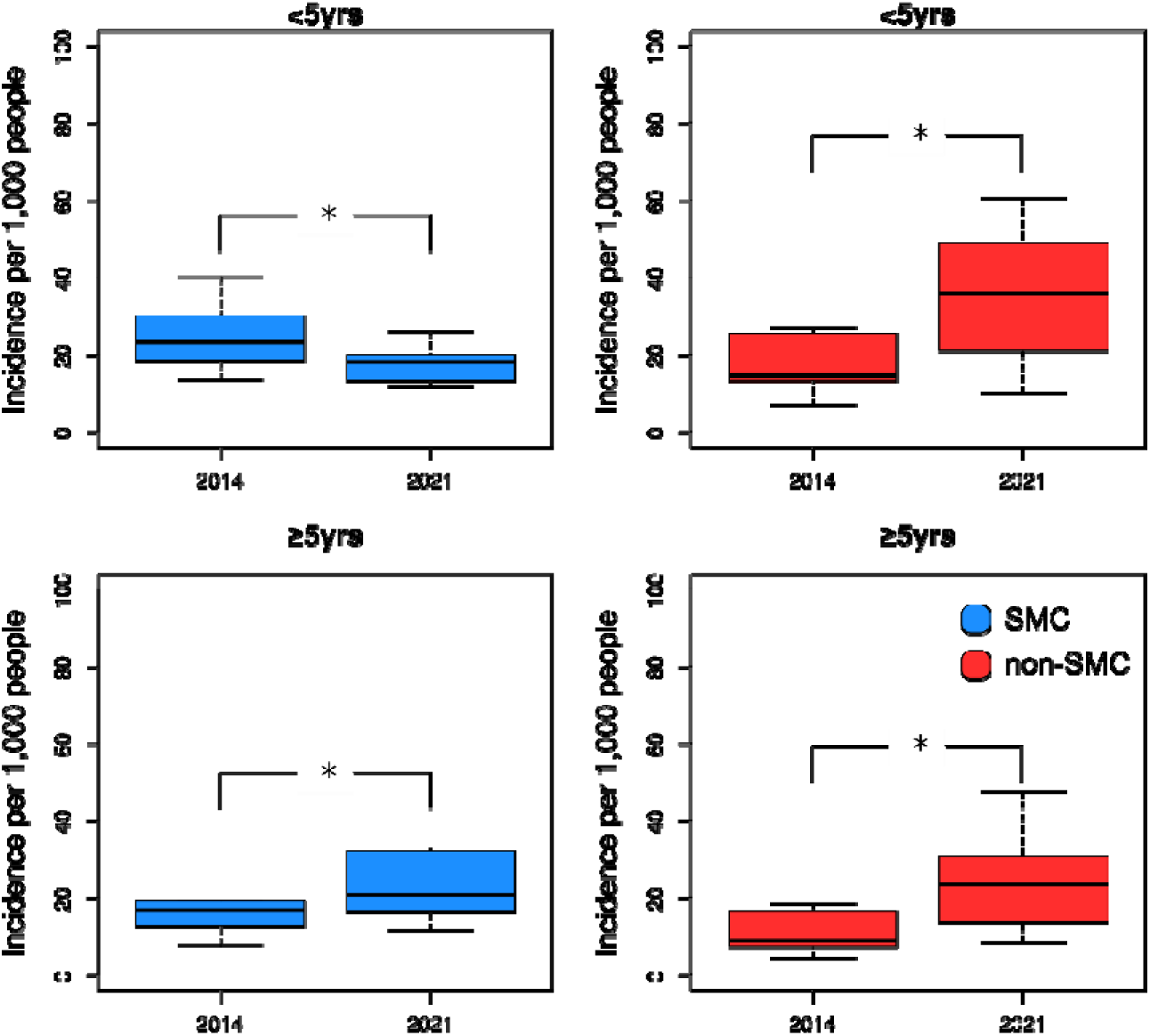
Boxplot comparing malaria incidence of SMC (left column) and non-SMC (right column) districts per age group of 2014 to April 2019 for <5 yrs age group and ≥5 yrs age group. The ‘*’ symbol represents a significant difference (p<0.05, Wilcoxon’s test) between SMC and non-SMC districts.

**Figure 10.**
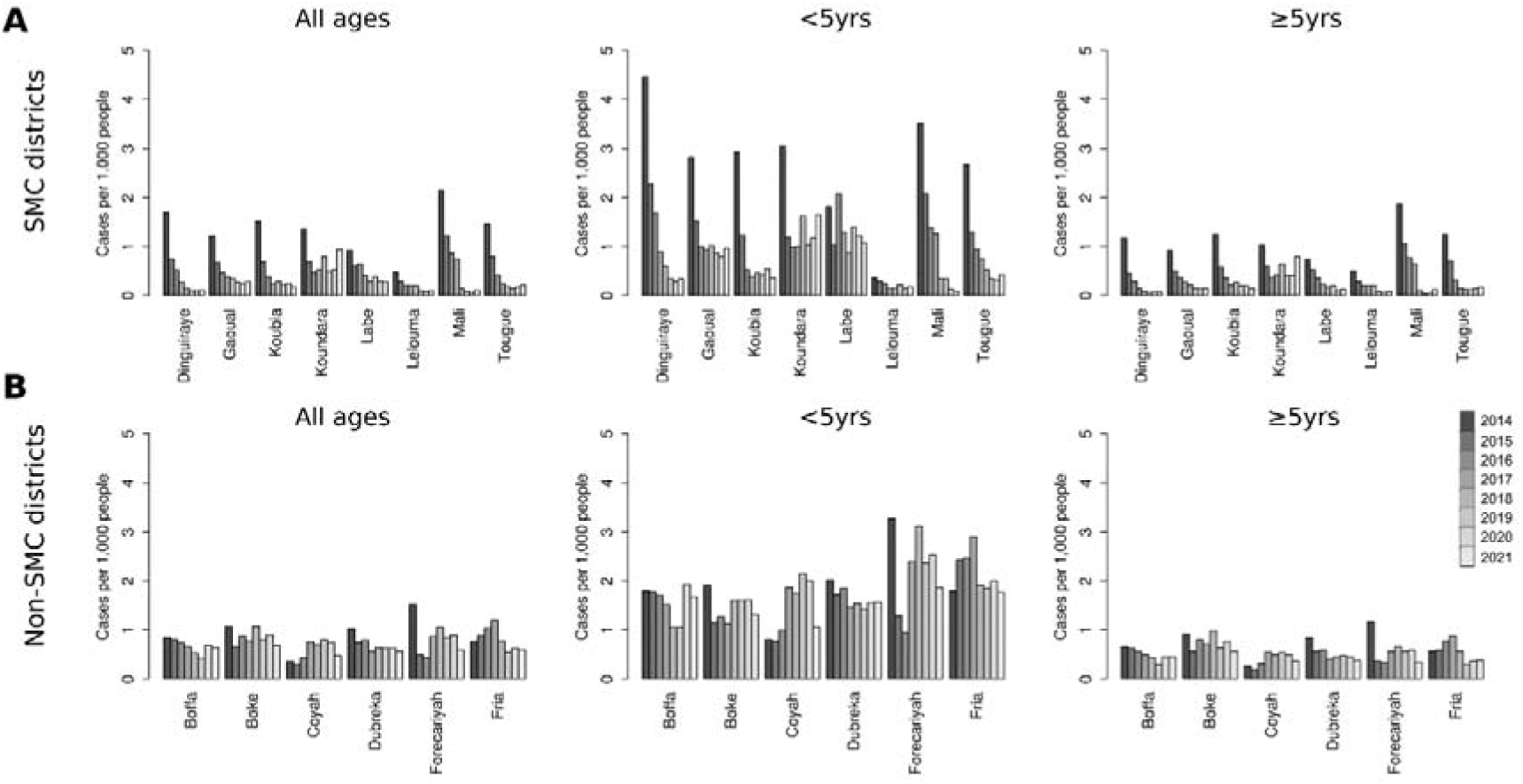
Incidence of severe malaria cases in SMC (A) and non-SMC (B) districts per age group and year. Plots show severe malaria incidence for all-age groups, <5yrs age group, and ≥5yrs age group in SMC (A) and non-SMC districts (B)

**Figure 11.**
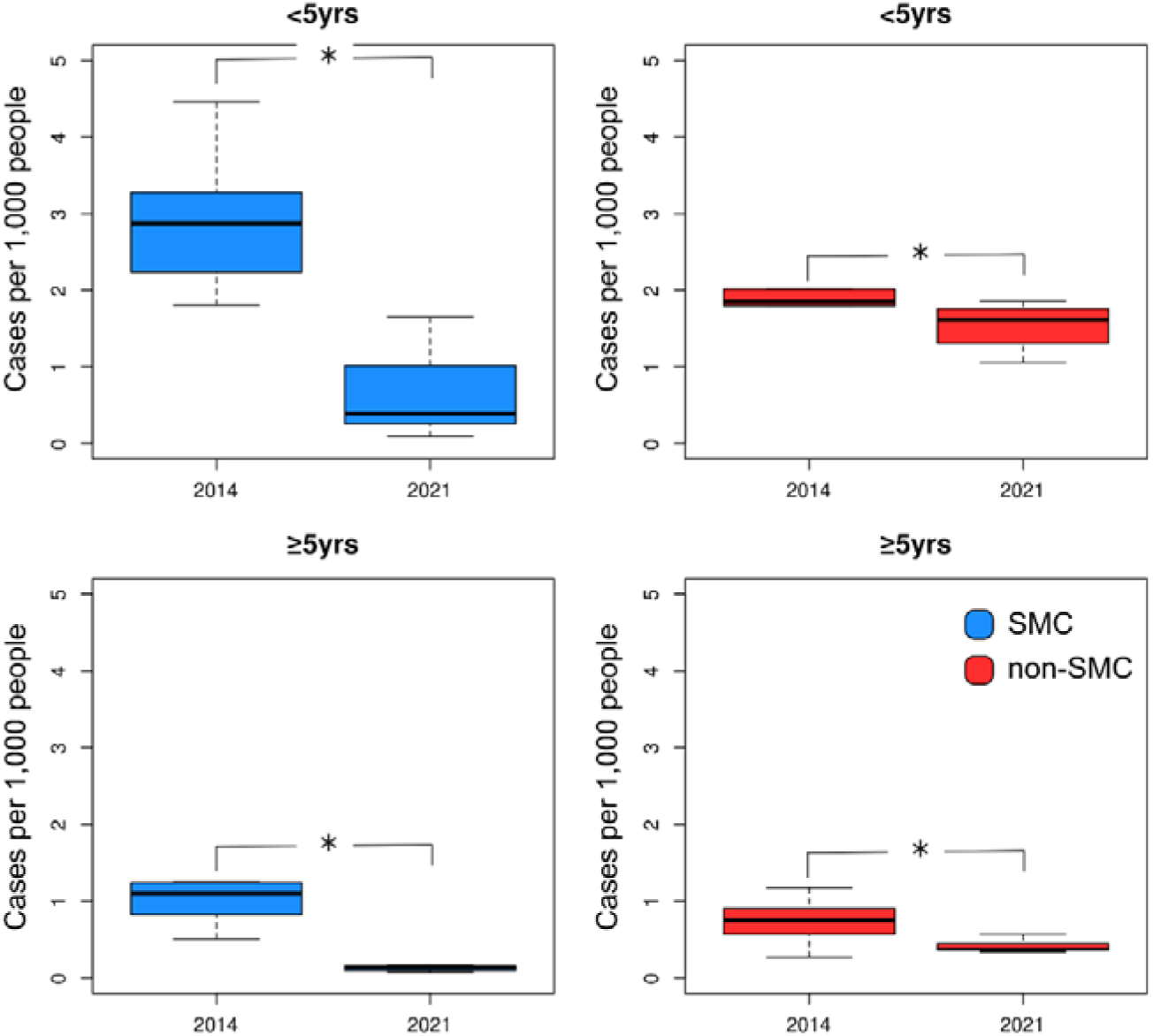
Boxplot comparing severe malaria cases per 1,000 people in SMC (left column) and non-SMC (right column) districts per age group of 2014 to April 2019 <5 yrs age group and ≥5 yrs age group. The ‘*’ symbol represents a significant difference (p<0.05, Wilcoxon’s test) between SMC district and non-SMC districts.

## DISCUSSION

Randomized controlled trials have shown that SMC can reduce the incidence of clinical malaria, parasitemia, anemia, severe malaria, and all-cause mortality in CU5, when effectively deployed.^4–7^ Several studies have analyzed the effectiveness of SMC when implemented at large operational scales, showing SMC protective effectiveness against clinical malaria ranging from 73–98% in CU5 at 28 and 42-days post-SMC administration, respectively.^28, 29^ However, these studies relied on large samples of at-risk populations (i.e., CU5), either following case / control study designs,^28, 29^ or using nationally representative surveys such as Demographic and Health Surveys or Malaria Indicator Surveys.^30, 31^ While such approaches are certainly robust, they can be onerous, time-consuming, and costly, and may not allow for continuous monitoring of the intervention across all SMC implementation areas.

As countries’ HMIS are strengthened, there is the opportunity to increasingly use data that is routinely (passively) collected by health facilities and reported by districts.^32–36^ Using district-level data from Guinea’s HMIS collected between 2014–2021, we show the programmatic impact of SMC in targeted districts, confirming the effectiveness of SMC shown in various controlled research studies. Our analyses show that—in a context of strengthened case management and medium to high coverage of LLINs—SMC significantly reduced the TPR, uncomplicated malaria incidence, and the incidence of severe malaria in CU5 in Guinea between 2014–2021. These findings also corroborate other recent analyses estimating SMC effectiveness through use of routine health information system data. Thus, in Chad, using generalized additive modelling, Richardson et al.^37^ estimated that SMC reduced the number of suspected and confirmed malaria in CU5 by 18% (95% CI: 6–28) and 19% (95% CI: 7–29) at primary health facilities in 23 health districts during the months of SMC implementation, respectively. Similarly, an evaluation of the ACCESS-SMC program using a difference-in-differences approach of DHIS2 data observed a 45.0% and 55.2% reduction in malaria outpatients after two years of SMC implementation in Burkina Faso and The Gambia, respectively; the study also reported substantial reductions in the number of malaria inpatients and malaria hospital deaths following SMC.^29^

Interestingly, we show that the effect of SMC is particularly pronounced in the dry season rather than the rainy season. We hypothesize that SMC results in CU5 either being fully protected from infection or, if infected, only having infections with low parasitemia (which may be due to incomplete adherence to the 3-day regimen, or drug resistance).^38^ Combined with the lower abundance and density of mosquitoes during the dry season, this then results in lower transmission rates, which leads to a lower incidence. Similar observations have been made in studies evaluating vector control interventions.^39^ These observations do show, however, that in Guinea SMC should possibly start earlier than in July (i.e., to have a greater effect on infection protection and transmission intensity during peak transmission season), and that studies monitoring SMC impact should potentially monitor effectiveness beyond the currently recommended 28–42 days post SMC administration.^40^

Moreover, unlike prior controlled academic research and other studies, our analyses also demonstrate an effect of SMC administered to CU5, albeit lower, on PO5, specifically for TPR, uncomplicated malaria incidence, and severe malaria incidence. Such effect seems plausible, given that—even in a context of decreasing malaria prevalence such as Guinea^1^—CU5 still represent a large proportion of all cases (i.e., 55.2% for the 2014–2021 study period; **Table 1**) and, thus, a substantial proportion of the human reservoir infecting anopheline mosquitoes. Any significant intervention effect reducing the CU5 *Plasmodium* reservoir will lower transmission intensity and, thus, spill over and reduce malariometric indicators in PO5.

As support continues for malaria elimination and additional means for malaria control are introduced (e.g., vaccines), alternative methods for measuring impact must be explored outside trial settings as controlled and observational research studies to investigate effectiveness at scale are very costly. Multiple impact evaluations and approaches that address gaps or biases in data and triangulating between data sources strengthens the plausibility of program impact. We used the CGAR as an analytical method. The CAGR is a commonly used methodology in the field of economics to describe changes in (economic) growth from the beginning to the end of a timeseries. The advantage of using CAGR compared to other analytical approaches—such as average annualized rates—is that the resulting estimate is not affected by fluctuations within the time series that could produce misleading results. The CAGR methodology has previously been used in the public health field to evaluate health care systems and effects of interventions through time.^41–43^ An additional advantage of the approach is that, because it is algebra-based, it does not require advanced data analytics or modelling expertise, and thus can be applied by program personnel with a basic understanding of calculus (e.g., in MS Excel worksheets, or even DHIS2).

Because SMC is consistently proving to be a high-coverage and effective intervention, several countries with seasonal malaria transmission outside of the Sahel are now piloting SMC to be included in their malaria intervention package (e.g., Uganda^44^ and Mozambique^45^). WHO recently amended the SMC guidelines to give countries flexibility to change the SMC regimen (e.g., by adding additional SMC cycles to cover longer peak transmission seasons, or by including older age groups).^8^ Such step is likely going to increase the effect of SMC even further, either in CU5 (if additional SMC cycles are added) or in older age groups (if older age groups are covered with SMC)—indeed recent studies in Burkina Faso, Mali and Senegal seem to corroborate this.^46–48^ Moreover, there is increased interest to use SMC campaigns to integrate or leverage other health interventions,^49–51^ the rationale being that (1) other health interventions that may not have as high population acceptance or coverage as SMC would benefit from the high acceptance and coverage of SMC; and (2) cost-efficiencies would be obtained, since combining intervention planning, implementation and monitoring would result in various economies of scale (e.g., training of health workers, transport costs, limiting health workers’ time to implement campaigns rather than providing health services).

### Limitations

Several potential caveats of our analyses should be highlighted. First, our analyses did not adjust for intervention coverage, either for SMC or other malaria interventions such as LLINs. Thus, we assume that programmatic coverage, use of, and adherence to these interventions is relatively homogenous and that the only difference in overall malaria intervention coverage between the two sets of districts included in our analyses is the administration of SMC to CU5. Post-SMC round assessments consistently showed programmatic coverage of 82–93% across districts included in our analyses (**Table 1**). Moreover, as per 2021 Malaria Indicator Survey data, LLIN use in CU5 in households with at least 1 LLINs was 60.4% in survey enumeration areas in SMC districts, compared to 65.6% for non-SMC districts.

Second, using routine HMIS data for impact evaluations can be problematic due to internal validity, completeness, and potential bias in estimates of effect, and caution must be exercised in interpreting analytical findings.^32^ Additionally, challenges with HMIS data are that they are dependent on the proportion of infected individual cases seeking treatment for malaria at public sector health facilities; the proportion of patients seeking care who are parasitologically diagnosed; and facilities reporting all suspected and confirmed malaria cases consistently over time. Thus, for example, even though SMC was implemented in Guinea since 2015, an increase in the number of fevers was observed in CU5, suggesting a lack of impact of SMC. However, during this period, changes in access to health services occurred, such as improvement in malaria case detection and diagnosis at facility and community levels, removal of user access fees for CU5 and pregnant women, and improvement in the availability of malaria diagnostic and case management commodities. We controlled for such bias by excluding health facilities with incomplete reporting (i.e., facilities with 10 months of missing data throughout the study period, and no more than 3 continuously missing months per year), and note that these facilities were evenly spread across SMC and non-SMC districts. We also note that our analyses of the SMC effect in CU5 was consistently observed across all malariometric indicators included in our analysed.

Third, malaria testing to identify cases in Guinea is largely RDT-based. Although used tests have a high sensitivity and specificity, it is possible that low parasitemic and asymptomatic infections were missed. It is unlikely, however, that such infections would have clustered in a specific spatiotemporal pattern across SMC compared to non-SMC districts, and, thus, would unlikely change our findings and conclusions.

Fourth, we delineated our analyses to district boundaries, rather than a defined area size (e.g., 25 km^2^). This was done because routinely collected HMIS data gets aggregated at that level and districts are the lowest administrative unit in Guinea that plan, implement and monitor malaria programming; any response to an increase in malaria cases would occur at district level.

Finally, spatial analyses performed using arbitrary spatial divisions such as district administrative boundaries can be affected by the modifiable areal unit problem.^52, 53^ For example, cases that were reported at a health facility in a given district could stem from households in a different district, and therefore, bias district-level malaria HMIS data. Thus, the possibility remains that cases got infected outside of their district (e.g., during travel).

## CONCLUSION

Our study provides evidence to support that—even in an operational programming context—adding SMC to the comprehensive package of malaria interventions yields a positive epidemiological impact and results in a greater reduction in TPR, malaria incidence and hospitalizations in CU5 than when implementing an intervention package without SMC. We also show that the use of routine HMIS data represents a viable method for assessing the epidemiological impact of public health interventions in the absence of trial studies, household surveys, and extensive covariate data. Further studies on the effectiveness of SMC at scale should use data obtained from a wider range of sources.

## Data Availability

All data produced in the present study are available upon reasonable request to the authors

## Acknowledgements

We thank Guinea’s Ministry of Health staff who participated in SMC programmatic efforts, as well as the communities in the study districts for their collaboration.

## Patient and public involvement

Patients and/or the public were not involved in the design, or conduct, or reporting, or dissemination plans of this research.

